# Exploring domains, clinical implications and environmental associations of a deep learning marker of biological ageing

**DOI:** 10.1101/2021.01.22.21250338

**Authors:** Alessandro Gialluisi, Augusto Di Castelnuovo, Simona Costanzo, Marialaura Bonaccio, Mariarosaria Persichillo, Sara Magnacca, Amalia De Curtis, Chiara Cerletti, Maria Benedetta Donati, Giovanni de Gaetano, Enrico Capobianco, Licia Iacoviello, on behalf of the Moli-sani Study Investigators

## Abstract

Deep Neural Networks (DNN) have been recently developed for the estimation of Biological Age (BA), the hypothetical underlying age of an organism, which can differ from its chronological age (CA). Although promising, these population-specific algorithms warrant further characterization and validation, since their biological, clinical and environmental correlates remain largely unexplored.

Here, an accurate DNN was trained to compute BA based on 36 circulating biomarkers in an Italian population (N=23,858; age≥35 years; 51.7% women). This estimate was heavily influenced by markers of metabolic, heart, kidney and liver function. The resulting Δage (BA-CA) significantly predicted mortality and hospitalization risk for all and specific causes. Slowed biological aging (Δage<0) was associated with higher physical and mental wellbeing, healthy lifestyles (e.g. adherence to Mediterranean diet) and higher socioeconomic status (educational attainment, household income and occupational status), while accelerated aging (Δage>0) was associated with smoking and obesity. Together, lifestyles and socioeconomic variables explained □48% of the total variance in Δage, potentially suggesting the existence of a genetic basis.

These findings validate blood-based biological aging as a marker of public health in adult Italians and provide a robust body of knowledge on its biological architecture, clinical implications and potential environmental influences.

## Introduction

Ageing is a time-dependent biological process characterized by functional decline[1], often implying cognitive impairment and depressive symptoms, as well as an increased risk of several chronic disorders[2]. However, ageing progression differs among individuals, with some people showing a slower decline than others[3, 4]. To investigate the reasons for these discrepancies among subjects and monitoring healthy ageing in the general population, different approaches have been recently proposed, based on the computation of Biological Age (BA), *i*.*e*. the hypothetical underlying age of an organism, which can differ from its chronological age (CA)[5, 6]. The latter approaches include methods based on blood biomarkers[7–9], which are conceived as markers of organismal BA, and methods based on organ-specific measures, such as spirometry[10] and structural neuroimaging[4]. Until now, BA estimates were mostly based on linear regression methods, in which BA is a function of one or few bodily measures[7]. However, machine learning (ML) algorithms have been recently developed[8, 9, 11], showing very good accuracy in predicting CA[6]. Among ML methods, a cost-effective model based on Deep Neural Networks (DNN, *i*.*e*. an algorithm which resembles the structure and functioning of the human brain) has been proposed to estimate BA, using blood biomarkers as input features and CA as label[8, 9, 12]. The resulting discrepancy between biological and chronological age (Δage) can assume either positive or negative values, indicating accelerated or decelerated biological ageing, respectively. Δage was reported to significantly predict mortality risk in North-American populations[8], suggesting it may represent an effective marker of public health. However, BA estimates are highly population-specific[8]. Moreover, although there has been progress in improving the interpretability of such models[13], very little is known on the environmental influences on Δage. At present, deep learning blood-based Δage results only associated with smoking status[12], as are more classical markers of biological age based on the Klemera-Doubal method[14]. Recently, another blood-based ageing clock was developed, MORTAL-bioage, based on the prediction of mortality risk by blood markers and chronological age through Cox Proportional Hazards (PH) models, and re-calibration of the risk in years[15]. The resulting biological age acceleration was negatively associated with physical activity, and positively with TV watching, and heavy alcohol drinking (>5 drinks/day), in addition to smoking status and number of cigarettes/day[15]. However, this clock had a different construction paradigm aimed at detecting mortality risk with the highest accuracy, and explicitly included age as predictor. We are not aware of any previous analysis of blood age with reference to socioeconomic indicators, while for other biological ageing (e.g. DNA methylation) clocks, significant associations were observed between accelerated ageing and disadvantaged childhood[16], as well as with years of education and economic disadvantage in adulthood[17].

Overall, there is a need to apply and validate deep learning based BA in different populations, testing their link with clinical outcomes and environmental influences, which remain largely unexplored. To this purpose, we developed a BA estimation algorithm based on blood biomarkers in an Italian adult population cohort, the Moli-sani study[6], and tested its predictive capacity of mortality and hospitalization risks for all and specific causes, as well as their association with common measures of mental and physical wellbeing. To identify environmental influences on BA, we also tested associations with lifestyle and socioeconomic factors, which are among the main modifiable risk factors of many health conditions[17, 18]. Moreover, there is a urgent need to identify the biological domains affected by biological ageing, so to target these domains for the development of future potential anti-ageing therapies. The wealth of clinical, biometric and environmental variables assessed within the Moli-sani study provides an unprecedented opportunity to investigate these aspects in a comprehensive manner, and to substantially contribute to the knowledge on such an easy-to-measure and financially efficient marker of ageing like blood age.

## Methods

### Population of study

All analyses were carried out within the Moli-sani study, a large population-based cohort of adult Italians (N=24,325; ≥35 years; 48.11% men) living in Molise, a small region located in central Italy with about 300,000 citizens. Between March 2005 and April 2010, men and women aged ≥35 years were randomly recruited from city-hall registries. Exclusion criteria were pregnancy, disturbances in understanding/willing processes, ongoing poly traumas or coma. The Moli-sani study was approved by the Ethical Committee of the Catholic University of Rome, and all the participants provided a written informed consent.

### Circulating biomarkers

A number of circulating biomarkers were tested within the Moli-sani cohort, including:

- lipid biomarkers, like total cholesterol, triglycerides, high (HDL) and low density lipoprotein (LDL), lipoprotein a (Lp(a)), apolipoprotein A1 (Apo-A1) and B (Apo-B);
- markers of glucose metabolism: glucose, C-peptide, insulin;
- liver enzymes: aspartate transaminase (AST) and alanine aminotransferase (ALT);
- cardiac and vascular markers: N-Terminal Pro-B-Type Natriuretic Peptide (NT-proBNP) and high-sensitivity cardiac troponin I;
- other hormones: testosterone and vitamin D;
- hemostasis markers: D-Dimer, fibrinogen;
- renal markers: uric acid, albumin, creatinine, cystatin-C;
- inflammation marker: high sensitivity C-reactive protein (CRP);
- common haemochrome markers including red blood cell count (RBC) and distribution width (RDW), hematocrit (Hct), hemoglobin levels (Hgb), mean corpuscular volume (MCV), mean corpuscular hemoglobin (MCH) and concentration (MCHC), total white blood cells (WBC), lymphocytes (LY), monocytes (MO), granulocytes (GR), neutrophils (NE), basophils (BAS) and eosinophils (EO); platelet count (Plt), mean platelet volume (MPV), platelet distribution width (PDW) and plateletcrit (PCT).

Details on laboratory measurements are reported in Supplementary Methods (Table S1).

### Data treatment and quality control

All the analyses were carried out in R[19]. Unreliable blood markers levels - i.e. leukocyte counts whose fractions summed <99% (86) or >101% (9) - were set to missing. Missing values (Table S1) were then imputed through both the k-nearest neighbor algorithm of the *VIM* package[20], (*kNN()* function, k=10), and the minimum/maximum imputation, the latter employed when an explicit missing code for indicating values below/above detection levels was available (for creatinine, Apo-A1, Apo-B, Lp(a), NT-proBNP, testosterone, troponin-I and VitD). Composite variables like total cholesterol, plateletcrit, hematocrit and MCH were removed to avoid collinearity, while we retained WBC and MCHC since these did not show very high correlations with other white and red cell parameters, respectively (Pearson’s r < 0.9; Figure S1). Participants reporting non-Italian ancestry (332) and/or non-faster status at the time of blood draw (135) were removed before analysis. After quality control, we had 23,858 participants (12,346 women; mean (SD) age = 55.9 (12.0) years) and 36 circulating markers available for analysis, which underwent min-max normalization.

### Deep Neural Network for computation of Biological Age

The *Keras* package (see URLs) was used to build a Deep Neural Network (DNN) algorithm for the prediction of BA, with circulating biomarkers, recruiting center and sex as input features, and CA of each participant as a label. DNN showed the best performance among different algorithms in a similar setting, i.e., with reference to the pioneering works on the topic[8, 9]. Similarly, we applied the best-performing combination of hyperparameter settings used in other studies[8, 9, 12], which implied five layers with 2000-1500-1000-500-1 neurons, dropout rate of 35% after each layer for regularization purposes, Mean Squared Error (MSE) as optimization loss function and AdaGrad as optimizer[21]. We trained the network over 1,000 epochs, in a 80% random extraction of the dataset (N=19,086), with a 20% internal validation set and a batch-size of 32, to avoid overfitting. Then we evaluated the accuracy of the algorithm in an independent test set (remaining 20% of the dataset, N=4,772), in terms of Mean Average Error (MAE), Pearson’s correlation (r) and Nagelke’s coefficient of determination (R^2^) in univariate linear regression between BA and CA. A comparison between the training and the test set is reported in Table S2, indicating no major differences in sociodemographic, lifestyles and clinical characteristics between the two subsets.

As in the indicated studies[8, 9, 12], we performed a permutation feature importance (PFI) analysis to identify those markers showing the largest influence on the prediction of BA. This implies shuffling measures of one marker at a time and then comparing the loss function (MSE) of the perturbed model with that of the full model (i.e. with no permuted feature). This analysis was carried out through the *explain()* and the *variable_importance()* functions of the DALEX package[22].

### Associations with clinical risk and healthy aging parameters

Once BA for each participant and the resulting discrepancy with CA were computed (Δage = BA - CA), we validated this new variable as public health marker in the test set (N=4,772). Information on lifestyles, wellbeing and socio-economic status had been collected at baseline[23] and classified as described below and elsewhere[24–26]. Passive follow-up data on clinical events of interest (deaths and hospitalizations) were collected and validated until December 31^st^ 2015, through linkage with hospital discharge records and regional mortality registry. A detailed description of variables and clinical events analyzed here can be found in Supplementary Methods.

First, we tested predictivity of mortality and first hospitalization incident risks, for all and specific causes, through multivariable Cox PH models, after checking all the basic assumptions (proportionality of hazards, linearity of effect and absence of influential observations). Cox PH regressions were performed both on the continuous Δage, and comparing participants with accelerated (Δage > 5 years, i.e. BA >> CA) and slowed ageing (Δage < −5 years, BA << CA), versus a reference class with BA ∼ CA (−5 ≤ Δage ≤ 5). Specific causes included cardiovascular (CVD), ischemic heart (IHD), cerebrovascular disease (CeVD) and cancer for both deaths and hospitalizations; deaths for other causes (including type-2 diabetes); and hospitalizations for type-2 diabetes. Survival analyses were initially adjusted for sex and CA (Model 1), to compare them with prominent field studies[5, 8]. Then, more conservative models were built, incrementally adjusted for i) main prevalent health conditions (CVD, cancer and diabetes; Model 2), and ii) lifestyles and socioeconomic (SES) variables (Model 3). Specifically, only those covariates showing at least a trend of association (p<0.2) with both incident mortality/hospitalizations risk and Δage were tested in the enriched survival models, namely all prevalent conditions, all lifestyles except for physical activity and all SES variables (see below). All the above mentioned covariates showed also significant associations with incident clinical risks (p<0.05), in line with previous findings made in the Moli-sani study[18, 25–31]. For this analysis, a conservative Bonferroni correction for multiple testing of two different types of clinical events – deaths and hospitalizations - was applied (α = 0.025).

Second, we tested associations of Δage with physical and mental wellbeing, as assessed through the validated Italian version of the self-administered Short Form 36 (SF-36) test[32]. This scale - often taken as a measure of healthy aging[6] - tests health-related quality of life (QoL) involving both physical and mental domains, which include physical functioning, role limitations due to physical health problems, bodily pain, general health perceptions, vitality, social functioning, role limitations due to emotional problems and mental health[33]. Again, incrementally adjusted models were applied, as above, and significance threshold was corrected for two different SF-36 subscales tested (α = 0.025).

### Associations with lifestyles and socioeconomic factors

Last, we tested associations of ΔAge with modifiable risk factors, including both lifestyles and socioeconomic variables. Among lifestyles, we analyzed smoking habits (current and ex-smokers vs non-smokers, number of cigarettes/day and years of smoking within current smokers); adherence to Mediterranean Diet (moderate and high vs low)[28, 34]; alcohol consumption (lifetime abstainers, former drinkers, current drinkers of 12.1-24, 24.1–48 and >48 g/day vs occasional or current drinkers of 1-12 g/day), as in [26]; and level of leisure time physical activity (MET-h/day)[35]. As a proxy of lifestyles, we analyzed obesity, both comparing participants with body mass index (BMI) 25-30 Kg/m^2^ and ≥30 Kg/m^2^ vs <25 Kg/m^2^, and testing continuous associations with Relative Fat Mass (RFM)[36].

Among socioeconomic variables, we tested associations with education level completed (lower, upper and post-secondary vs none/primary), occupational class (unemployed/unclassified, retired/housewives, manual skilled, unskilled and non-manual skilled vs professional/managerial workers), yearly household income (>40k, 25-40k, 10-25k vs <10k Euros/year), childhood SES (computed as in [18] and classified as high, intermediate and upper-low vs low), and housing status (dwelling one or more ownerships vs living in a rented flat).

These factors were first analyzed separately in linear models adjusted for CA and sex (taking ΔAge as outcome), then in a multivariable setting including all the variables showing an association p<0.1 with ΔAge in univariate models (see *Results* below). To avoid potential overfitting bias due to multicollinearity of exposures, we applied a stepwise regression approach based on Akaike Information Criterion (AIC), through the *stepAIC()* function of the MASS package in R[37] (see Supplementary Methods). For this analysis, we applied a Bonferroni correction for ten independent variables – five lifestyles and five SES indicators – resulting in α = 5×10^−3^.

## Results

### Accuracy of the DNN algorithm and most predictive biological features

The best performing DNN showed accurate BA estimates, compared to CA values (MAE = 6.00 years; r = 0.76; R^2^ = 0.57; Figure S2), in line with previous studies in the field[8, 13]. Based on the PFI analysis, the most important biological features influencing BA values in our DNN were cystatin-C, NT-proBNP and sex, all showing an increase in the loss (MSE) function of the algorithm >30%, compared to the baseline model (Figure 1). The resulting difference between BA and CA (Δage) showed slightly higher values in men compared to women (mean (SD): −0.61 (7.63) *vs* −1.19 (7.91); Student’s t = −2.55, p = 0.01). Δage was normally distributed (Figure S3) and negatively correlated with CA in the test set (Figure S4), as seen for other biological ageing acceleration estimators[16, 38, 39]).

**Figure 1.**
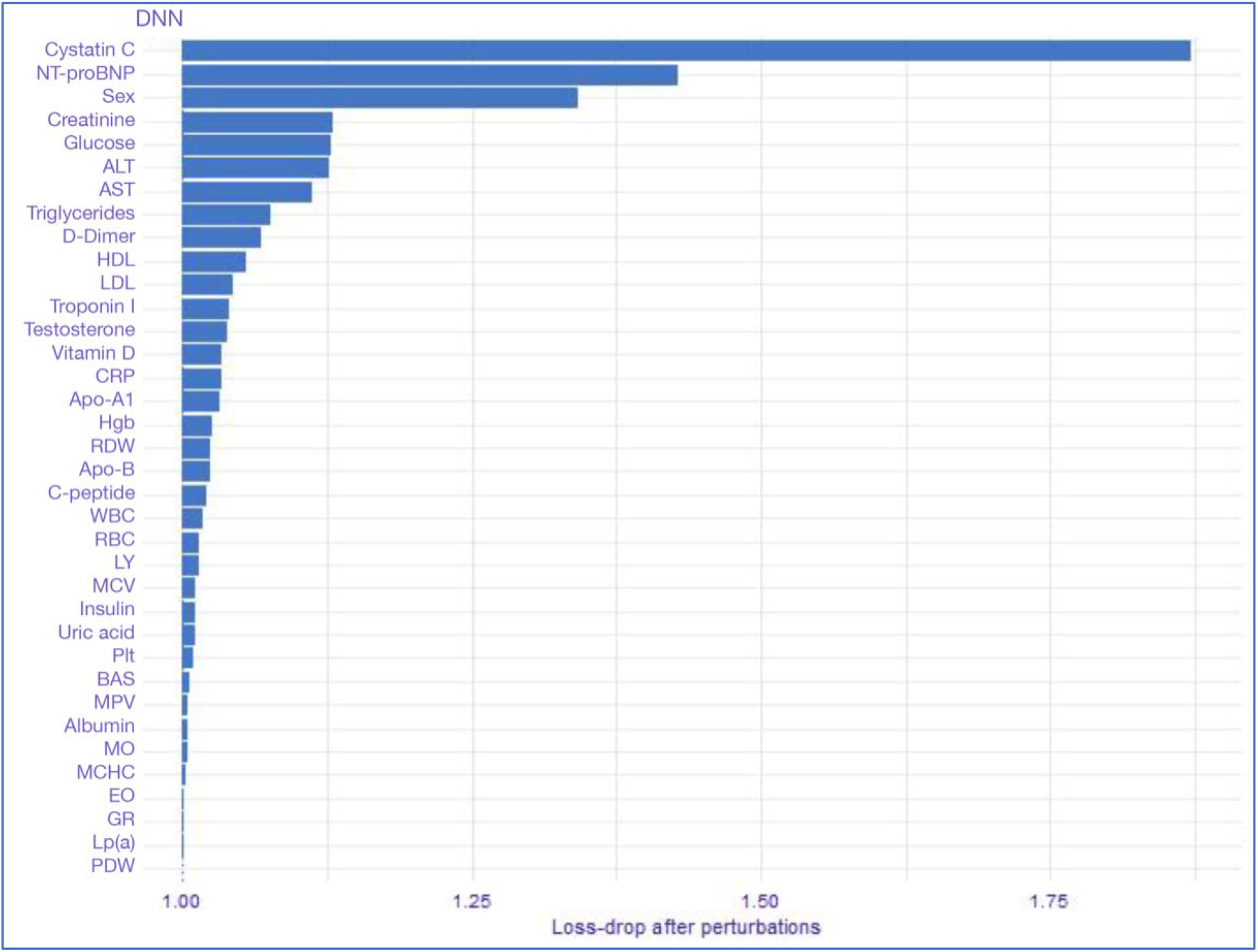
Permutation Feature Importance (PFI) analysis. Bars indicate the importance of each biological feature used for the prediction of BA in the DNN algorithm, based on the ratio between loss function (MSE) in the perturbed model (i.e. after permutation of a given variable) and loss function in the full model (with no permuted variable). The higher the ratio, the more the perturbed model is altered and the more important is the permuted feature. Abbreviations: NT-proBNP = NT-proB-type Natriuretic Peptide; ALT = alanine aminotransferase; AST = aspartate transaminase; HDL/LDL = high/low density lipoprotein; Lp(a) = lipoprotein-a; Apo-A1/-B = apolipoprotein A1/B; VitD = vitamin D; CRP = high sensitivity C-reactive protein; RBC = red blood cell count; RDW = red cell distribution width; Hgb = hemoglobin; MCHC = mean corpuscular hemoglobin concentration; LY, MO, GR, NE, BAS, EO, WBC = lymphocyte, monocyte, granulocyte, neutrophil, basophil, eosinophil and total white blood cell count; Plt = platelet count; MPV = mean platelet volume; PDW = platelet distribution width.

### Clinical significance of Δage

Δage significantly predicted the incident risk of all-cause mortality and first hospitalizations (Table 1a, b and Figure 2a, b). Over 4,767 participants with mortality data available in the test set (283 death events, median follow-up 8.27 years), each yearly increase in Δage was associated with a 7% (CI=5-10%) increase in all-cause mortality risk (p=5.8×10^−11^; Table 1a). Subjects with BA >> CA showed a 134% (49-266%) increase of risk, while participants with BA << CA showed a 49% (32- 61%) decrease, compared to those with BA falling within CA ± 5 years. This trend was substantially consistent across different causes of death, except for cancer, and in more conservative models adjusted for prevalent health conditions (Model 2), lifestyles and socioeconomic variables associated with Δage (Model 3; Table 1a).

**Table 1.** Associations of Δage with incident a) mortality and b) first hospitalization risk.

| Cause of death | N (events) | Model 1<br>HR [95% CI]<br>(p-value) | Model 2<br>HR [95% CI]<br>(p-value) | Model 3<br>HR [95% CI]<br>(p-value) |
| --- | --- | --- | --- | --- |
| **All causes** | **4,767 (283)** | **1.07 [1.05; 1.10]**<br>($5.8 \times 10^{-11}$) | **1.06 [1.04; 1.09]**<br>($8.5 \times 10^{-9}$) | **1.06 [1.04; 1.08]**<br>($6.5 \times 10^{-7}$) |
| **CVD** | **4,765 (100)** | **1.13 [1.09; 1.17]**<br>($6.6 \times 10^{-10}$) | **1.12 [1.08; 1.16]**<br>($9.8 \times 10^{-9}$) | **1.11 [1.07; 1.16]**<br>($2.9 \times 10^{-7}$) |
| **IHD + CeVD** | **4,765 (60)** | **1.11 [1.06; 1.17]**<br>($7.4 \times 10^{-6}$) | **1.10 [1.05; 1.16]**<br>($3.6 \times 10^{-5}$) | **1.10 [1.05; 1.16]**<br>($1.1 \times 10^{-4}$) |
| Cancer<sup>a</sup> | 4,563 (83)<sup>a</sup> | 1.02 [0.98; 1.06]<br>(0.27) | 1.02 [0.98; 1.06]<br>(0.39) | 1.01 [0.97; 1.05]<br>(0.59) |
| **Other** | **4,767 (71)** | **1.08 [1.04; 1.13]**<br>($2.2 \times 10^{-4}$) | **1.08 [1.03; 1.12]**<br>($7.1 \times 10^{-4}$) | **1.08 [1.04; 1.12]**<br>($2.4 \times 10^{-5}$) |

**b)**
| Cause of hospitalization | N (events) | Model 1<br>HR [95% CI]<br>(p-value) | Model 2<br>HR [95% CI]<br>(p-value) | Model 3<br>HR [95% CI]<br>(p-value) |
| --- | --- | --- | --- | --- |
| **All causes** | **4,771 (2,027)** | **1.03 [1.02; 1.04]**<br>($9.1 \times 10^{-16}$) | **1.03 [1.02; 1.03]**<br>($2.2 \times 10^{-11}$) | **1.02 [1.01; 1.03]**<br>($2.0 \times 10^{-7}$) |
| **CVD** | **4,771 (771)** | **1.05 [1.04; 1.06]**<br>($3.8 \times 10^{-14}$) | **1.04 [1.03; 1.05]**<br>($1.6 \times 10^{-9}$) | **1.04 [1.02; 1.05]**<br>($1.6 \times 10^{-7}$) |
| **IHD** | **4,771 (187)** | **1.05 [1.03; 1.08]**<br>($2.8 \times 10^{-5}$) | 1.03 [1.00; 1.06]<br>(0.02) | 1.02 [1.00; 1.05]<br>(0.08) |
| CeVD | 4,771 (135) | 1.03 [1.00; 1.06]<br>(0.07) | 1.02 [0.99; 1.05]<br>(0.13) | 1.02 [0.98; 1.05]<br>(0.35) |
| Cancer<sup>a</sup> | 4,569 (235)<sup>a</sup> | 0.99 [0.97; 1.01]<br>(0.26) | 0.99 [0.96; 1.01]<br>(0.21) | 0.98 [0.96; 1.01]<br>(0.19) |
| **Diabetes** | **4,771 (80)** | **1.09 [1.05; 1.13]**<br>($5.4 \times 10^{-6}$) | **1.06 [1.02; 1.10]**<br>($5.4 \times 10^{-3}$) | 1.04 [1.00; 1.09]<br>(0.05) |

Risk estimates for a) deaths and b) hospitalizations for all and specific causes are expressed as Hazard Ratios (HR) with 95% Confidence Interval (CI) per year increase in Δage, as calculated by Cox PH regression models with time-on-study on the time scale and competing risk of dying/being hospitalized for other causes, adjusted for sex and Chronological Age (CA). Statistically significant Hazard Ratios ( $p < 0.025$ ) are highlighted in bold. Final sample size (N) indicates the number of samples actually analyzed with a case-complete approach in the test set (Model 1), after removing samples with missing event and follow-up data. <sup>a</sup> Participants with a diagnosis of cancer at recruitment or with an incident cancer event in the first year of follow-up were removed for this specific analysis, as in [61]. Abbreviations: CVD = cardiovascular disease; CeVD = cerebrovascular disease; IHD = ischemic heart disease.

**Figure 2.**
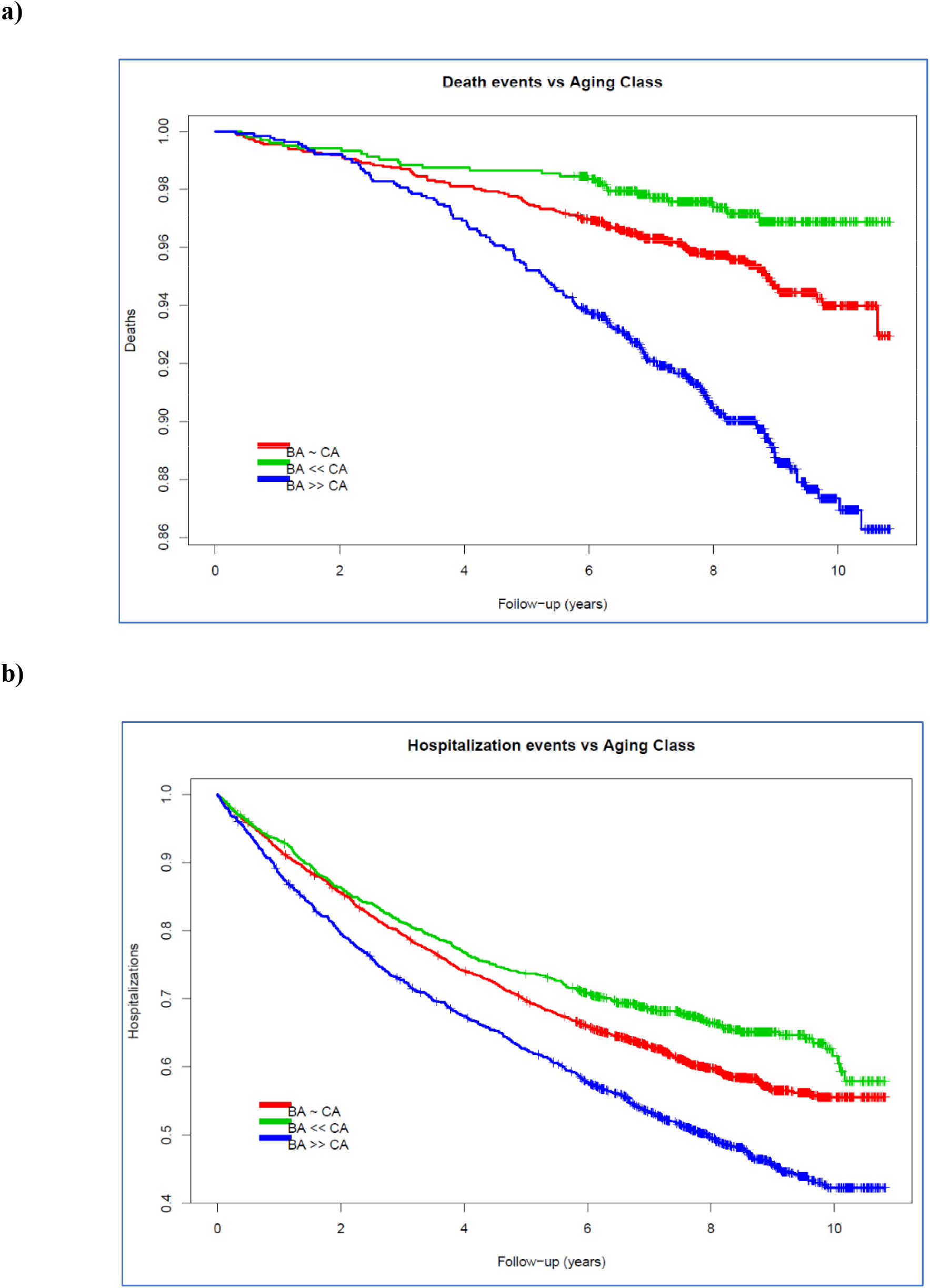
Survival curves of all-cause a) mortality and b) first hospitalization risk vs Δge. Subjects with predicted Biological Age (BA) within five years from their Chronological Age (CA) (red curve) were compared to participants with BA at least five years younger (green curve) and older than their CA (blue curve), as in [8].

Similarly, the analysis of first hospitalizations (N=4,771, 2,027 all-cause admissions, median follow-up 7.10 years) revealed an increased risk by 3% (CI=2-4%; p=9.1×10^−16^) per year increase in Δage (p=9.1×10^−16^; Table 1b), with biologically older participants showing a 30% (14-48%) increase and younger participants showing a 27% (29-35%) decrease of risk compared to those with BA ∼ CA. Again, this trend was confirmed by hospitalizations for all specific causes but cancer and remained significant in enriched models adjusting for prevalent chronic conditions and lifestyles/SES, for all but IHD events (Table 1b).

Both physical and mental wellbeing showed a negative association with Δage (β(SE) = −0.30(0.09) and −0.81(0.10) years per SD increase in SF-36 scales). However, incremental adjustments for health conditions and main determinants of disease notably reduced effect sizes of these associations (Table S3).

### Association of Δage with main lifestyle and socioeconomic factors

The analysis of modifiable risk factors revealed statistically significant associations of Δ*age* with lifestyle variables or their proxies (Table 2a). Δage was significantly associated with smoking status (β(SE) = 1.38(0.22) years for current vs never-smokers), adherence to Mediterranean Diet (β(SE) = −0.90(0.30) for high vs low adherence), and alcohol consumption (β(SE) = 0.92(0.23) years for life- time abstainers vs occasional and current drinkers who drank <12 g/day). Obesity, which is heavily influenced by lifestyles, was positively associated with Δage (β(SE) = 1.73(0.22) years for participants with BMI ≥30 vs <25 Kg/m^2^), as was RFM too, but not leisure time physical activity (Table 2b).

**Table 2.** Association of Δage with main a) categorical and b) continuous lifestyle variables or proxies.

| Lifestyle variable | Levels/Variables | Beta | SE | p | N |
| --- | --- | --- | --- | --- | --- |
| **Smoking status**<br>(Missing = 10) | Never smokers<br>(reference) | - | - | - | 2390 |
|  | **Current smokers** | **1.38** | **0.22** | **$2.1 \times 10^{-10}$** | **1066** |
|  | Previous smokers | 0.34 | 0.21 | 0.11 | 1306 |
| **Obesity**<br>(Missing = 6) | <25 Kg/m<sup>2</sup><br>(reference) | - | - | - | 1260 |
|  | [25-30[ Kg/m<sup>2</sup> | 0.13 | 0.21 | 0.52 | 1997 |
|  | **$\geq 30$ Kg/m<sup>2</sup>** | **1.73** | **0.22** | **$4.6 \times 10^{-15}$** | **1509** |
| **Adherence to Mediterranean Diet**<br>(Trichopoulou score, Missing = 22) | Low (0-3) | - | - | - | 1485 |
|  | Moderate (4-6) | -0.47 | 0.18 | 0.01 | 2788 |
|  | **High (7-9)** | **-0.90** | **0.30** | **$2.8 \times 10^{-3}$** | **477** |
| **Alcohol consumption**<br>(Missing = 188) | Occasional/current drinkers<br>(1–12 g/day)<br>(reference) | - | - | - | 1202 |
|  | **Life-time abstainers** | **0.92** | **0.23** | **$9.4 \times 10^{-5}$** | **1221** |
|  | Former drinkers | 0.03 | 0.42 | 0.94 | 215 |
|  | Current drinkers (24.1-48 g/day) | 0.05 | 0.27 | 0.87 | 806 |
|  | Current drinkers (12.1-24 g/day) | 0.15 | 0.28 | 0.58 | 682 |
|  | Current drinkers (>48 g/day) | -0.20 | 0.33 | 0.56 | 458 |

**b)**
| Lifestyle variable | Unit | Beta | SE | p | N |
| --- | --- | --- | --- | --- | --- |
| Smoking habits<br>(within Current smokers) | Cigarettes/day | 0.02 | 0.01 | 0.24 | 1066 |
|  | Years of smoking | 0.04 | 0.03 | 0.13 | 1066 |
| Leisure physical activity | MET-h/day | -0.001 | 0.08 | 0.99 | 4772 |
| **Obesity** | **Relative Fat Mass** | **1.02** | **0.15** | **$4.6 \times 10^{-11}$** | **4766** |

All associations were adjusted for sex and Chronological Age (CA). Beta values indicate the variation in Δage (years) **a)** associated with each class compared to reference (for categorical variables) and **b)** per Standard Deviation (SD) increase in the continuous variables. Statistically significant associations ( $p < 5 \times 10^{-3}$ ) are highlighted in bold.

Among socioeconomic variables (Table 3), educational attainment showed a significant trend of association across all the levels tested, but only upper secondary (β(SE) = 1.17(0.24) years, compared to primary school) and post-secondary level (1.24(0.32) years) survived correction for multiple testing (p < 5×10^−3^). A concordant trend was observed for occupational class, with unemployed presenting the highest increase in Δage compared to professional/managerial workers (2.86(0.70) years). Likewise, yearly household income was consistently associated with Δage (β(SE) = −1.92(0.41) years for participants who declared >40k vs <10k Euros), as was housing status, whose association however did not survive correction for multiple testing.

**Table 3.** Association of Δage (BA-CA) with socioeconomic factors.

| SES variable | Levels | Beta | SE | p | N |
| --- | --- | --- | --- | --- | --- |
| Education level completed<br>Missing: 7 | None/primary school (reference) | - | - | - | 1236 |
|  | Lower secondary | -0.60 | 0.25 | 0.02 | 1342 |
|  | **Upper secondary** | **-1.17** | **0.24** | **$1.96 \times 10^{-6}$** | **1632** |
|  | **Post-secondary** | **-1.24** | **0.32** | **$8.29 \times 10^{-5}$** | **555** |
| Occupational class<br>Missing: 0 | Professional and managerial (reference) | - | - | - | 963 |
|  | Non-manual skilled | 0.31 | 0.23 | 0.18 | 1664 |
|  | **Manual unskilled** | **1.01** | **0.27** | **$1.7 \times 10^{-4}$** | **844** |
|  | **Manual skilled** | **1.31** | **0.26** | **$5.6 \times 10^{-7}$** | **982** |
|  | **Retired/housewife** | **1.44** | **0.42** | **$6.1 \times 10^{-4}$** | **248** |
|  | **Unemployed/unclassified** | **2.86** | **0.7** | **$4.8 \times 10^{-5}$** | **71** |
| Yearly household income (Euros)<br>Missing: 0 | <10.000 (reference) | - | - | - | 318 |
|  | [10.000-25.000] | -0.12 | 0.36 | 0.74 | 1402 |
|  | **]25.000-40.000]** | **-1.15** | **0.37** | **$1.9 \times 10^{-3}$** | **1010** |
|  | **>40.000** | **-1.92** | **0.41** | **$2.2 \times 10^{-6}$** | **552** |
|  | Not declared | -0.48 | 0.35 | 0.17 | 1490 |
| SES during childhood<br>Missing: 142 | Low (reference) | - | - | - | 657 |
|  | Upper-low | -0.12 | 0.26 | 0.65 | 1640 |
|  | Intermediate | -0.28 | 0.28 | 0.32 | 1423 |
|  | High | -0.29 | 0.32 | 0.36 | 910 |
| Housing<br>Missing: 8 | Rented flat (reference) | - | - | - | 454 |
|  | Dwelling ownership | -0.07 | 0.28 | 0.81 | 3884 |
| | >1 dwelling ownership | -1.02 | 0.39 | $8.3 \times 10^{-3}$ | 426 |

All associations were adjusted for sex and Chronological Age (CA). Beta values indicate the variation in Δage (years) associated with each class, compared to reference. Statistically significant associations ( $p < 5 \times 10^{-3}$ ) are highlighted in bold.

In a stepwise multivariable regression modelling together lifestyles and SES factors, smoking status, obesity, adherence to Mediterranean Diet, alcohol consumption, occupational class, household income and housing status were retained (adjusted-R^2^ = 0.48). Although all the variables showed concordant associations with univariate models, only those with smoking status, obesity and occupational class remained statistically significant, while others were only nominally or marginally significant (Table S4).

## Discussion

We reported the first application of a deep learning algorithm to estimate biological age based on blood markers (BA) in an Italian population. This provided not only biological hints into the ageing process, but also a detailed characterization of the resulting accelerated ageing index (Δage), from a biological, clinical and epidemiological perspective.

### Biological domains of blood ageing

Among the wealth of biological features used in the DNN, cystatin-C was by far the most influent in the estimation of BA, followed by NT-proBNP and sex. Cystatin-C is an index of glomerular filtration rate and of kidney function, along with creatinine[40], which was also among the most influential markers. A lower cystatin-C level has been associated with successful ageing (i.e. free of cardiovascular disease, cancer, and chronic obstructive pulmonary disease, with intact physical and cognitive functioning) in elders, even within a range of relatively normal kidney function[41]. Interestingly, DNA methylation-based surrogates for cystatin-C significantly predict mortality risk and have been integrated (among others) in a recent version of the epigenetic clock, GrimAge[42]. NT-proBNP is a precursor of the Brain Natriuretic Peptide (BNP), which is produced by the left heart ventricle under wall stretching stress and is an index of conditions like heart failure, left ventricular dysfunction, acute coronary syndromes and stroke[43]. Sex has been reported as one of the most predictive features of BA also in previous studies along with other prominent features such as glucose, liver enzymes and cholesterol markers, all having an impact on ageing and lifespan[8, 9, 12, 13]. This evidence suggests that the domains with the largest influence on the biological ageing process include metabolic, heart, kidney and liver function.

### Clinical implications and correlates of blood ageing

When characterizing the discrepancy between biological and chronological age from a clinical point of view, we observed that Δage significantly predicted incident all-cause mortality risk in our population, with effect sizes similar to those detected in North-American populations[8, 15]. Here, we extended this evidence to specific causes of mortality like CVD – in line with previous evidence for other ageing clocks[44] - and to hospitalization risk for all and specific causes like CVD and diabetes, albeit in this case the associated risk was generally more modest. To our knowledge, this is the first time evidence is provided that links biological ageing and hospitalizations, although an association between subjective age (i.e. how old one person feels) and incident hospitalizations was previously reported in US cohorts[45]. Of note, we did not observe any significant association of accelerated biological ageing with incident cancer risk, which was instead previously observed for a mixed blood-lung measure[14]. Similarly DNA methylation age was previously reported to increase incident risk of colorectal[46] and breast cancer[47]. At present, it is difficult to say whether this is due to the low number of heterogenous cancer cases analyzed (which unavoidably affects power), or to the fact that blood age as computed here actually does not represent a risk factor for cancer, contrary to other aging clocks like methylation[48]. Larger studies focusing on blood-based BA and on specific cancer types will allow to clarify this aspect.

Δage showed significant negative associations with health-related quality of life scales representing physical and mental wellbeing, which are considered important components of healthy ageing[3] and have been associated with perceived ageing[49]. Although these scales have never been tested with a pure blood age clock, QoL is associated with frailty[50] and the SF-36 physical component correlates positively with accelerated biological ageing based on mixed instrumental (blood and respiratory) biomarkers[51]. Importantly, by modelling the physical and mental component in the same regression, our results support the existence of independent associations of the two QoL domains tested, specifically underlining the importance of the mental component in decelerating ageing, in addition to the physical one.

Of interest, while adjustment for main chronic conditions did not heavily affect the associations observed with incident mortality/hospitalization risk and only partly attenuated those with physical/mental wellbeing, adjustment for lifestyles and SES showed different behaviors. As for mortality risk, this did not determine any substantial change in estimation, while this was more evident for hospitalizations, especially for diabetes and mostly IHD events. The associations with QoL subscales were notably affected by adjustment for the main determinants of disease, with mental wellbeing becoming non-significant after correcting for lifestyles and SES factors. These observations may be explained by three alternative hypotheses. First, lifestyles/SES may represent confounders in the relationship between biological aging and clinical outcomes, especially for hospitalization risk and measures of frailty like physical/mental QoL. Second, biological aging discrepancy may represent a mediator in the associations between lifestyles/SES and clinical outcomes, reported elsewhere[18, 25–31]. While this represent a fascinating and previously untested hypothesis, the data currently available do not allow to formally test it (see *Strengths and Limitations* below). Third, the associations surviving corrections for lifestyles/SES suggest that these may be partly due to common genetic influences between biological aging and clinical risks/indices. In our view, it is conceivable that all these explanations may apply.

### Environmental associations with blood ageing

Disentangling the relationship between Δage and the main determinants of disease, namely lifestyles and socioeconomic status[17], we corroborated previous associations with smoking status[12, 15] and provided evidence that adherence to a healthy (Mediterranean) diet slows down ageing, in line with previous observational[17, 52] and interventional studies analyzing epigenetic age[53]. On the contrary, the lack of evidence of a link between physical activity and slowed biological ageing is discordant with previous evidence reported for blood[15] and brain age[54], but concordant with studies on DNA methylation age[17, 52]. Similarly, a protective effect of moderate alcohol drinking on ageing was apparent in occasional/current drinkers who drank 1–12 g/day, in comparison with life-time abstainers. Our finding is not directly comparable with previous reports of increased blood ageing for subjects drinking >2 and >5 alcohol units/day (equivalent to >24 and >60 g/day, respectively) compared to abstainers[15], but it is in line with a protective effect of moderate alcohol consumption against mortality and hospitalization risk[26, 55]. A proxy measure of lifestyles, obesity, was also positively associated with ageing acceleration, considering both BMI-based weight categories and linear associations with RFM. This is in line with previous associations with other mixed blood-lung [14] and epigenetic clocks[17, 52].

The analysis of socioeconomic variables suggested a potential slowing-down effect of higher SES, with more educated participants being biologically younger than their CA compared to less educated ones. Similarly, participants in higher financial income strata were biologically younger than those in the lowest, as were workers in with higher occupational classes compared to unemployed. Although to our knowledge no BA estimator based on blood markers has ever been tested with reference to SES, our findings are consistent with inverse associations of years of education with slowed biological ageing, both at the brain[54] and at the epigenetic level[17]. The latter DNA methylation measure was also positively associated with an economic disadvantage index based on low income, education, unemployment, occupational class, and other variables[17]. Conversely, we found no evidence of association between childhood SES and Δage, which was instead observed elsewhere for a DNA methylation clock[16]. This discrepancy may be explained by different populations tested and methods of SES assessment or, more likely, by the different BA estimator used, leading to hypothesize that childhood disadvantage may affect the epigenetic domain rather than circulating markers. Further studies are needed to clarify this aspect, as well as whether the “pro-ageing” effect of SES is reversible, *e*.*g*. through the use of trajectories[17].

The associations above were generally confirmed in a multivariable stepwise regression model, although important variables like education level and healthy (MeDi) diet were not retained or not significant anymore. This could be justified by the type of analysis carried out, where collinear variables not explaining additional variance in the model were removed through the stepwise approach, or did not explain a significant proportion of Δage variance. In this perspective, it may well be that, e.g. part of the association with MeDi was also explained by obesity – which is heavily influenced by diet[56] – or that the effect of education was fully explained by other correlated SES variables[29, 57]. Since this represent the first multivariable analysis of lifestyles and SES determinants of biological ageing, there are no terms of comparison and further studies are needed to clarify these overlaps. Prominently, all the lifestyles/SES indicators tested explained □48% of the total Δage variance, supporting again the existence of genetic underpinnings to account for at least part of the remaining variance, which would be hardly explained in toto by other non-genetic factors like environmental pollution. Although this hypothesis is in line with previous evidence for brain age[11, 58, 59], the genetic basis of blood ageing remain totally unexplored.

### Strengths and Limitations

This study represents one of the largest and probably broadest assessment of clinical implications and environmental influences of blood age. Some limitations, however, should be mentioned. First, we did not perform a systematic hyperparameter tuning due to the need of extra powered computational infrastructure, therefore limiting our explorations to hyperparameter settings computed as optimal by previous studies [8, 9, 12]. However, the lower sample size of our study and the accuracy of our algorithm in line with previous ones, indicate only a small chance for significant performance improvement of our DNN. Second, other biological age markers showed a higher accuracy than blood age, *e*.*g* brain age[5, 11], although these are based on more expensive and less commonly tested neuroimaging measures that have limited applicability to large cohorts and in public health programs. Third, other biological age acceleration markers are better in predicting mortality risk[15], but only focus on accurately estimating this risk, which is only one aspect of the ageing process. Other aspects that should not remain neglected include the relationship with equally important components, like hospitalizations and wellbeing, which we tested here for the first time. Last, the fact that blood markers, lifestyles/SES and QoL were assessed all at the same time point does not currently allow to further disentangle their complex relationship and to clarify the role of lifestyles and SES factors, e.g. whether they represent only confounders of the relationship between Δage and clinical outcomes, or whether they represent ancestral exposures, with Δage playing a mediation role. The upcoming availability of active follow-up data in the Moli-sani study will help clarifying this aspect.

### Conclusions

In conclusion, although the proposed deep learning blood-based BA marker needs further investigation to clarify clinical implications, environmental and genetic influences, our study provides sufficient proof of validity as a public health tool and calls for development and testing in different populations. Moreover, it corroborates the existence of a genetic basis for biological ageing. Last but not least, our data suggest the utilization of Δage as an index of personalized health in the general population, potentially driving public health interventions aimed at slowing down biological ageing.

## Data Availability

The data and codes supporting the findings of this study are available from the corresponding author and/or from the senior author of the manuscript upon request.

## Acknowledgements, Funding and Competing Interests

We thank the BiomarCaRE Investigators for testing some of the markers used in this study and Dr Nina Tirozzi for the refinement of artwork. This study was partially supported by the Italian Ministry of Economic Development (PLATONE project, bando “Agenda Digitale” PON I&C 2014- 2020; Prog. n. F/080032/01-03/X35) and by the Italian Ministry of Health (grant RF-2018- 12367074 to GdG and SC). AG, MB and SC were supported by Fondazione Umberto Veronesi. The Moli-sani Study Investigators thank the Associazione Cuore Sano Onlus (Campobasso, Italy) for its cultural and financial support. The enrolment phase of the Moli-sani Study was supported by research grants from Pfizer Foundation (Rome, Italy), the Italian Ministry of University and Research (MIUR, Rome, Italy) – Programma Triennale di Ricerca, Decreto no. 1588 and Instrumentation Laboratory, Milan, Italy. Funders had no role in this study design, collection, analysis, and interpretation of data, nor in the writing and submission phase of the manuscript. All Authors were and are independent from funders and declare no conflicts of interest.

## Declarations

### Authors’ Contributions

LI, MBD, ADiC, CC and GdG originally inspired the Moli-sani Study. AG and LI contributed to the conception and design of this study. MP, ADeC and SM carried out biological sample management and measurements, while SC and ADiC performed statistical data elaboration and curation in the Moli-sani Study. EC, MB, SC and ADiC provided technical and theoretical support for statistical analyses. AG analyzed the data and wrote the first draft of the manuscript, with contributions and critical review from all the co-authors.

### Ethics Approval and Consent to participate

The Moli-sani Study was approved by the ethical committee of the Catholic University of Rome (on March 8, 2004; approval nr: A-931/03-138-04/CE 2004) and all the participants provided written informed consent.

### Supplementary Files

Supplementary Methods and Results.

## URLs

R: https://www.r-project.org/

VIM package: https://cran.r-project.org/web/packages/VIM/index.html

Keras package: https://cran.r-project.org/web/packages/keras/index.html

DALEX package: https://cran.r-project.org/web/packages/DALEX/citation.html

MASS package: https://cran.r-project.org/web/packages/MASS/

