## Supplementary for "Exploring domains, clinical implications and environmental associations of a deep learning marker of biological ageing"

**Supplementary Methods**

***Circulating biomarkers***

Blood samples were obtained between 7 and 9 AM from participants who had fasted overnight and had refrained from smoking for at least 6 hours (h). Biochemical analyses were performed in the centralised Moli-sani laboratory, as described elsewhere[1]. Haemochromocytometric analyses were performed by cell counter (Coulter HMX, Beckman Coulter, Milan, Italy) within 3 h from venepuncture. Analyses of creatinine, apoliproteins A1/B, C-peptide, cystatin C, insulin, lipoprotein a, N-Terminal Pro-B-Type Natriuretic Peptide, testosterone, high sensitivity troponin I and vitamin D were performed within the BiomarCaRE Consortium and have been described in details elsewhere (see <https://cordis.europa.eu/project/id/278913/reporting/it>)[2]. Main information and descriptive statistics for all the circulating biomarkers currently available in the Moli-sani study is reported in Table S1.

**Table S1. Details and main descriptive statistics of all the blood markers currently available in the Moli-sani study.**

| Variable | Source | Unit | Observations | # Missing | Missing (%) | Min | Max | Median | Mean | SD |
| --- | --- | --- | --- | --- | --- | --- | --- | --- | --- | --- |
| LY | blood | % | 23450 | 875 | 3.6 | 6.30 | 90.20 | 32.40 | 32.64 | 7.33 |
| MO | blood | % | 23449 | 876 | 3.6 | 0.70 | 34.30 | 7.00 | 7.10 | 2.10 |
| GR | blood | % | 23447 | 878 | 3.6 | 5.00 | 90.60 | 60.40 | 60.27 | 7.75 |
| EO | blood | % | 17290 | 7035 | 28.9 | 0.10 | 33.70 | 2.20 | 2.62 | 1.75 |
| BA | blood | % | 16885 | 7440 | 30.6 | 0.02 | 19.90 | 0.60 | 0.62 | 0.40 |
| NE | blood | % | 17309 | 7016 | 28.8 | 2.11 | 88.30 | 56.80 | 56.67 | 8.14 |
| WBC | blood | ×10^3^/µl | 23666 | 659 | 2.7 | 1.40 | 68.80 | 6.00 | 6.23 | 1.78 |
| RBC | blood | ×10^6^/µl | 23666 | 659 | 2.7 | 2.67 | 498.00 | 4.89 | 4.94 | 3.24 |
| Hgb | blood | g/dl | 23664 | 661 | 2.7 | 6.60 | 24.10 | 14.50 | 14.49 | 1.51 |
| Hct | blood | % | 23666 | 659 | 2.7 | 13.50 | 67.90 | 43.30 | 43.28 | 4.03 |
| MCV | blood | fl | 23665 | 660 | 2.7 | 10.20 | 123.90 | 89.00 | 88.33 | 6.04 |
| MCH | blood | pg | 23665 | 660 | 2.7 | 14.10 | 42.20 | 29.90 | 29.58 | 2.45 |
| MCHC | blood | g/dl | 23666 | 659 | 2.7 | 22.50 | 317.00 | 33.60 | 33.47 | 2.17 |
| RDW | blood | % | 23544 | 781 | 3.2 | 0.27 | 33.90 | 12.70 | 12.93 | 1.23 |
| Plt | blood | ×10^3^/µl | 23666 | 659 | 2.7 | 2.33 | 1091.00 | 243.00 | 248.92 | 64.15 |
| MPV | blood | fl | 23401 | 924 | 3.8 | 5.20 | 16.40 | 8.80 | 9.01 | 1.23 |
| PCT | blood | % | 23388 | 937 | 3.9 | 0.02 | 1.11 | 0.22 | 0.22 | 0.06 |
| PDW | blood | % | 23252 | 1073 | 4.4 | 8.20 | 62.60 | 16.20 | 15.64 | 1.78 |
| Total Cholesterol | serum | mg/dl | 24174 | 151 | 0.6 | 55.00 | 604.00 | 211.00 | 213.18 | 41.77 |
| HDL | serum | mg/dl | 24172 | 153 | 0.6 | 14.00 | 135.00 | 56.00 | 57.53 | 14.89 |
| LDL | serum | mg/dl | 23818 | 507 | 2.1 | 3.60 | 416.40 | 128.20 | 130.12 | 35.10 |
| Triglycerides | serum | mg/dl | 24173 | 152 | 0.6 | 19.00 | 950.00 | 109.00 | 130.06 | 85.72 |
| Glucose | serum | mg/dl | 24174 | 151 | 0.6 | 12.00 | 443.00 | 97.00 | 101.49 | 25.40 |
| CRP | serum | mg/L | 24297 | 28 | 0.1 | 0.01 | 20.00 | 1.52 | 2.59 | 3.26 |
| D-dimer | plasma citrate | ng/ml | 21824 | 2501 | 10.3 | 2.15 | 1000.00 | 179.00 | 199.19 | 109.51 |
| Albumin | serum | gr/dl | 16270 | 8055 | 33.1 | 1.80 | 5.60 | 4.20 | 4.21 | 0.32 |
| ALT | serum | U/lt | 16270 | 8055 | 33.1 | 3.00 | 329.00 | 20.00 | 23.47 | 15.41 |
| AST | serum | U/lt | 16268 | 8057 | 33.1 | 11.00 | 316.00 | 23.00 | 24.61 | 10.34 |
| Uric acid | serum | mg/dl | 16266 | 8059 | 33.1 | 0.00 | 22.90 | 5.20 | 5.30 | 1.50 |
| Creatinine | serum | mg/dl | 23647 | 678 | 2.8 | 0.33 | 7.13 | 0.78 | 0.82 | 0.21 |
| Apo-A1 | serum | g/L | 23674 | 651 | 2.7 | 0.15 | 3.30 | 1.51 | 1.55 | 0.32 |
| Apo-B | serum | g/L | 23683 | 642 | 2.6 | 0.00 | 2.13 | 0.96 | 0.98 | 0.24 |
| C-peptide | serum | ng/ml | 23419 | 906 | 3.7 | 0.01 | 12.08 | 1.60 | 1.76 | 0.83 |
| Cystatin-C | serum | mg/L | 23666 | 659 | 2.7 | 0.01 | 9.34 | 0.95 | 0.99 | 0.25 |
| Insulin | serum | pmoli/L | 23504 | 821 | 3.4 | 2.30 | 1459.70 | 51.10 | 60.01 | 44.11 |
| Lp(a) | serum | mg/dl | 22999 | 1326 | 5.5 | 0.00 | 90.00 | 11.30 | 18.08 | 18.83 |
| NT-proBNP | EDTA plasma | pg/ml | 22453 | 1872 | 7.7 | 5.00 | 24396.00 | 50.18 | 98.85 | 283.00 |
| Testosterone | serum | nmoli/L | 22366 | 1959 | 8.1 | 0.45 | 35.00 | 1.96 | 9.17 | 9.40 |
| Troponin-I | serum | pg/ml | 23767 | 558 | 2.3 | 0.00 | 2538.90 | 2.20 | 3.76 | 24.19 |
| VitD | serum | ng/ml | 23452 | 873 | 3.6 | 0.00 | 138.30 | 17.50 | 18.91 | 9.26 |

Abbreviations: NT-proBNP = N-Terminal Pro-B-Type Natriuretic Peptide; ALT = alanine aminotransferase; AST = aspartate transaminase; HDL/LDL = high/low density lipoprotein; Lp(a) = lipoprotein-a; Apo-A1/-B = apolipoprotein A1/B; VitD = vitamin D; CRP = high sensitivity C-reactive protein; RBC = red blood cell count; RDW = red cell distribution width; Hct = hematocrit; Hgb = hemoglobin; MCH = mean corpuscular hemoglobin; MCHC = mean corpuscular hemoglobin concentration; LY, MO, GR, NE, BA EO, WBC = lymphocyte, monocyte, granulocyte, neutrophil, basophil, eosinophil and total white blood cell count; Plt = platelet count; MPV = mean platelet volume; PDW = platelet distribution width; PCT = plateletcrit.

***Exposures and covariates***

Physical and mental wellbeing were assessed through the validated Italian version of the self-administered SF-36 test (SF36-MCS)[3], a widely used and thoroughly validated scale assessing health-related quality of life (QoL)[4]. The questionnaire contains 36 items measuring eight multi-item parameters of health status covering the following domains: physical functioning, role limitations due to physical health problems, bodily pain, general health perceptions, vitality, social functioning, role limitations due to emotional problems and mental health, which include depression and anxiety components. The first four domains deal with physical aspects and generate the physical component score, while the other four reflect psychological features and generate the mental component score[4]. For each domain, a score ranking from 0 (worst health) to 100 (best health) was calculated as the weighted sum of the questions relevant to that domain. To obtain the global scores for each component (mental and physical), relevant domains are first transformed into a z-score (assigning equal weight to each item), then the resulting z-scores are combined into a global z-score through a weighted mean, using weights that resulted from a principal component analysis. Finally, the global scores are standardized to a normal distribution with mean = 50 and SD = 10[5].

For *smoking status*, subjects were assigned to three categories based on their cigarette smoking habits: *smokers, ex-smokers* (i.e. subjects who quitted at least one year before the interview) and *never-smokers* (reference class).

Leisure-time *physical activity* was assessed through a structured questionnaire and expressed as daily energy expenditure in metabolic equivalent task-hours (MET-h/day)[6].

Food intake was assessed through the validated Italian EPIC food frequency questionnaire[7]. Adherence to *Mediterranean Diet* was defined according to the Mediterranean Diet Score, ranging from 0 (low adherence) to 9 (high adherence)[8]. Then we defined three adherence classes: low (Trichopoulou score 0-3, used as reference), moderate (4-6) and high (7-9). The EPIC questionnaire also allowed to compute *alcohol consumption* habits along with additional questions, as described by [9]. Participants were classified based on the estimated daily alcohol volume consumed in the year before enrolment: life-time abstainers, former drinkers, occasional drinkers and current drinkers who drank 1–12 (reference class), 12.1–24, 24.1–48 and >48 g/day.

Height and weight were measured for each participant, as well as waist circumference (cm), which was measured in the middle between the 12^th^ rib and the iliac crest, while hip circumference (cm) was measured around the buttocks. Then *Body mass index (BMI)* was calculated, and participants were grouped into three categories: <25, 25–29.9, and ≥30 kg/m^2^. Since *Relative Fat Mass (RFM)* has been reported to be more accurate than BMI in estimating whole-body fat percentage measured by dual energy X-ray absorptiometry, and to improve body fat-defined obesity misclassification[10], we computed and analysed this parameter, computed as

*RFM = 64 − (20 × height/waist circumference) + (12 × sex),*

where height and waist are in the same scale unit, sex = 0 for men and 1 for women[10].

Socioeconomic status (SES) information was self-reported and/or collected by a structured questionnaire administered by trained personnel.

*Educational attainment* was defined as the education level completed and subjects were divided into four classes: None/Primary (reference), Lower secondary, Upper secondary and Post-secondar*y*.

Current *occupational social class* was classified based on the Registrar General’s occupation classification scheme, and ranked as previously described by[11]. Based on this rank, five occupational classes were defined: professional/managerial (reference), skilled non-manual, skilled manual, partly skilled/unskilled and unemployed/unclassified.

Self-reported *household income*, expressed as earned Euros per year, was defined as a five-level variable: <10,000; [10,000–25,000[; [25,000–40,000[; >40,000, and missing values, which were collapsed into a non-respondent category. The lowest yearly income class was taken as reference in the analyses.

Socioeconomic status at 8 years of age (*childhood SES*) was defined using retrospective questions on i) housing tenure (rented, 1 dwelling ownership, >1 dwelling ownership); ii) access to hot water; and iii) overcrowding in the household (i.e. the ratio between the number of rooms available and of persons living in the household)[12]. Each individual SES factor was coded as 0–1 to generate a total childhood SES score ranging from 0 to 3, which was classified as high (3), intermediate (2) upper-low (1) and low (0; reference).

*Housing tenure* was defined as for childhood SES, and classified into three categories: 1 dwelling ownership, >1 dwelling ownership and living in a rented flat, which was used as referent[13].

***Longitudinal data on hospitalizations and mortality***

Overall and cause-specific mortality updated to December 31^st^, 2015 was assessed through interrogation of the Italian mortality (ReNCaM) registry and linkage with the Moli-sani database through unique identifier codes for each participant. Death events were validated by Italian death certificates (ISTAT form) and coded according to the International Classification of Diseases, 9^th^ version (ICD-9; accessed on January 4, 2018). Cardiovascular and cancer mortality was defined when the underlying cause of death included ICD-9 codes 390-459 and 140-208, respectively[14]. Within cardiovascular mortality, ICD-9 codes 430-438 were used to define deaths for cerebrovascular disease (i.e. both haemorrhagic and ischemic stroke), and codes 410-414 and 429 for ischemic heart disease[14], then these were collapsed into a single mortality cause due to the paucity of events within each class[15]. Non-cardiovascular and non-cancer causes of death were included in death events for all causes, including mortality for type 2 diabetes (ICD-9 code 250), due to the paucity of events attributed to this cause in the analysed sub-cohort (6).

First hospital admissions were recorded by direct linkage with the Molise regional registry of hospital discharge records (HDR), updated to December 31^st^, 2015, as previously described[9]. Regional HDRs include hospitalizations of all citizens living in a given region, from both private and public hospitals, and present summary records for each hospital stay including patient demographics, clinical and administrative/financial information about the hospitalization. A hospitalization was defined as any stay lasting ≥ 24 hours in any hospital, clinic, emergency room or similar, except for diabetes (see below). If a patient was transferred to another hospital or facility, a single hospitalization was counted*.* Primary and secondary diagnoses for hospitalization were coded using ICD-9 and Major Diagnostic Categories (MDC) of the Italian Diagnosis Related Groups classification (version 24), and were elected as cause of hospitalization accordingly[9]. In the present study, we focused on hospitalization events for all causes, excluding pregnancy complications, childbirth, rehabilitation, chemotherapy and/or radiotherapy. Also, we analysed hospitalizations for specific causes, namely cardiovascular disease (defined as primary diagnosis MDC: 05, or ICD-9: 390-459 and MDC: 01/05), ischemic heart disease (primary diagnosis ICD-9: 410-414, or surgical procedure with ICD-9: 36.0-36.1), cerebrovascular events (primary diagnosis ICD-9: 430-434,436-438, or surgical procedure with ICD-9: 38.12), cancer (primary or first secondary diagnosis ICD-9: 140-172, 174-208; and MDC codes specific for malignant neoplasm procedures), and type 2 diabetes (primary diagnosis ICD-9: 250, which included day hospital). In the case of hospitalizations, diabetes was analysed separately thanks to the availability of a sufficient number of events (80).

**Figure S1. Correlation plot of the 36 blood markers tested and Chronological Age (CA) within the population of study (N=23,858).**

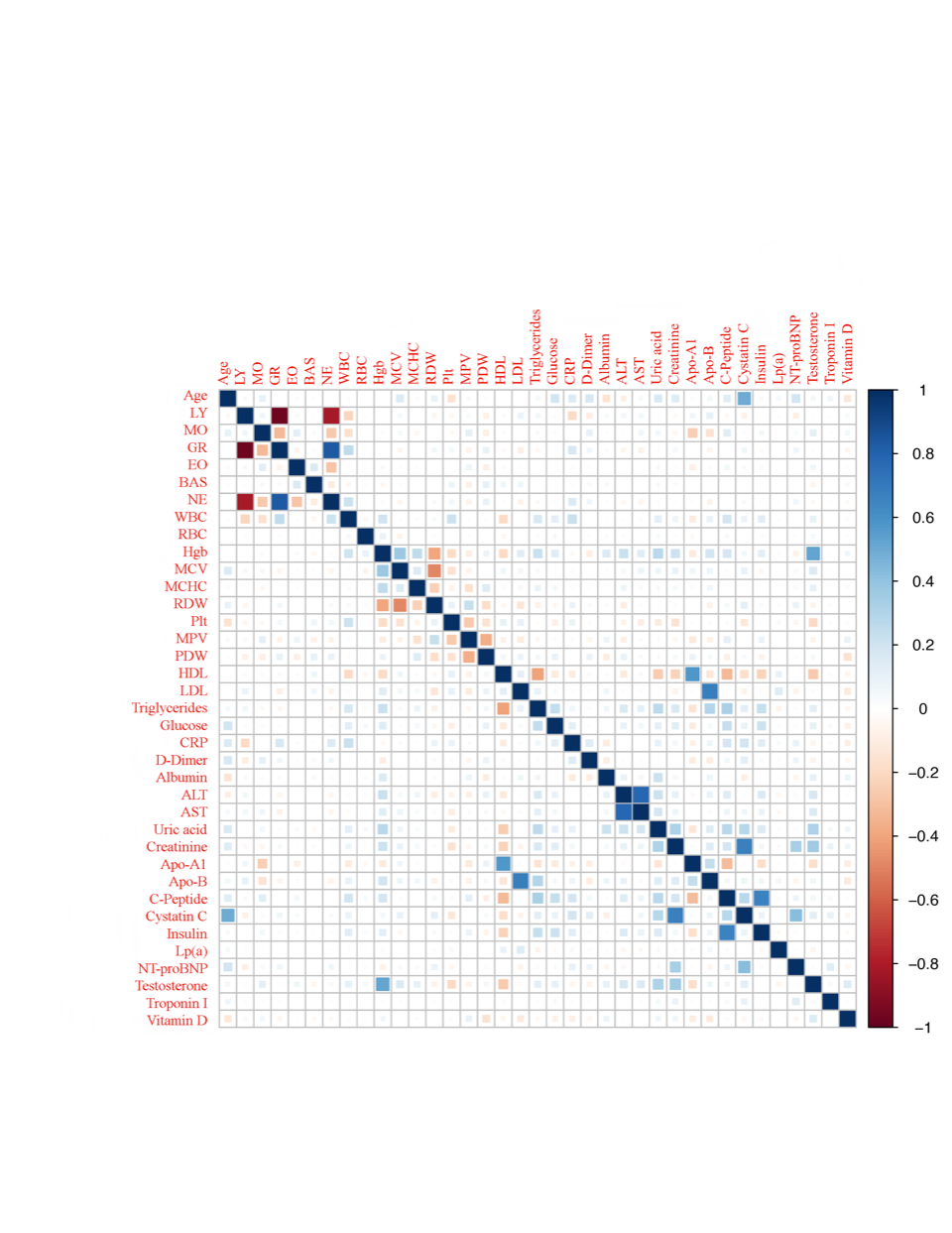

Pairwise Pearsons’s r correlation coefficients are displayed through color intensities (blue for positive and red for negative correlations). Abbreviations: NT-proBNP = N-Terminal Pro-B-Type Natriuretic Peptide; ALT = alanine aminotransferase; AST = aspartate transaminase; HDL/LDL = high/low density lipoprotein; Lp(a) = lipoprotein-a; Apo-A1/-B = apolipoprotein A1/B; VitD = vitamin D; CRP = high sensitivity C-reactive protein; RBC = red blood cell count; RDW = red cell distribution width; Hgb = hemoglobin; MCHC = mean corpuscular hemoglobin concentration; LY, MO, GR, NE, BAS, EO, WBC = lymphocyte, monocyte, granulocyte, neutrophil, basophil, eosinophil and total white blood cell count; Plt = platelet count; MPV = mean platelet volume; PDW = platelet distribution width.

**Table S2. Comparison between test set and training set, and characteristics of the whole Moli-sani cohort.**

| Variable | Test Set  (N=4,772) | Training Set  (N=19,086) | Moli-sani cohort  (N=24,325) | P-value  (test vs training) |
| --- | --- | --- | --- | --- |
| Sex (males) | 48.2 % | 48.3 % | 48.1 % | 0.91 |
| Age (years; mean, sd) | 55.9 (11.9) | 55.9 (12.0) | 55.8 (12.0) | 0.40 |
| *Education*  Primary  Lower secondary  Upper secondary  Post-secondary  Unknown | 25.9 %  28.1 %  34.2 %  11.6 %  0.2 % | 26.0 %  27.5 %  33.8 %  12.5 %  0.2 % | 25.8 %  27.7 %  33.9 %  12.4 %  0.2 % | 0.37 |
| *Health conditions*  CVD  Cancer  Diabetes | 6.1 %  3.2 %  4.9 % | 5.9 %  3.6 %  5.1 % | 5.9 %  3.5%  5.0% | 0.24  0.47  0.96 |
| *Lifestyle factors*  Current Smokers  Alcohol intake  (g/day; mean, sd)  Energy intake  (Kcal/day; mean, sd)  MeDi score (mean, sd)  Physical activity  (meth/day; mean, sd) | 22.3 %  15.9 (22.8)  2,082 (666)  4.4 (1.6)  3.5 (4.0) | 22.9 %  16.0 (22.6)  2,087 (669)  4.4 (1.6)  3.5 (4.0) | 22.9 %  16.0 (22.6)  2,079 (668)  4.3 (1.6)  3.5 (4.0) | 0.62  0.28  0.66  0.76  0.44 |
| *Obesity*  Obese participants  (BMI ≥ 30 Kg/m^2^)  RFM (mean, sd) | 31.6 %  35.6 (7.9) | 29.8 %  35.5 (7.8) | 30.0 %  35.5 (7.8) | 0.08  0.25 |

Since the whole Moli-sani cohort includes both the test and the training set and is largely overlapping with the latter, only p-values resulting from statistical comparison between the two (independent) subsets are reported. Chi-squared test was applied to education levels, CVD, cancer, obesity (BMI) and smoking status; Fisher Exact Test to sex and diabetes, unpaired t-test to age, MeDi score and calory intake; and Wilcoxon signed rank test to RFM, alcohol intake and physical activity levels (see Appendix for further details on the definition of these covariates). Abbreviations: CVD = cardiovascular disease; MeDi = adherence score to Mediterranean Diet (MeDi)[8]; BMI = body mass index; RFM = Relative Fat Mass[10]. Note: the Moli-sani cohort also includes 467 samples removed during QC (see main text).

***Multivariable stepwise regressions***

We applied multivariable regressions to model ΔAge as a function of all lifestyles and SES factors together, including all variables showing an association p < 0.1 with ΔAge in the baseline models (see *Results* in the main text). To avoid potential overfitting bias due to multicollinearity of exposures – which may lead to an inflation bias of Beta and R^2^ coefficients[16] – we used a stepwise regression approach based on Akaike Information Criterion (AIC). This was implemented through the *stepAIC()* function of the MASS package in R[17] (see URLs), using ΔAge ⁓ sex + CA as baseline model and keeping in the final model only those variables determining a decrease in the AIC both in forward and in backward stepwise regressions (“both” option).

AIC is defined as

***AIC = 2*k - 2 * LogLike (model),***

Where *k* is the number of parameters (independent variables) in the model and *LogLike* represents the natural logarithm of the likelihood of the model (the higher, the most predictive the model). AIC represents a tradeoff between the goodness-of-fit of a given model and its parsimony, i.e. the number of parameters included, and models with the lowest AIC are usually preferred. In other words, in this analysis only those independent variables which significantly contributed to an increase in the total variance explained by the model - in spite of the addition of a parameter to the regression – were kept, allowing to “clean” the models for potential bias introduced by other non-necessary variables. Indeed, this has been suggested as the preferential approach when many variables are treated as simultaneous exposures in a multivariable model[16].

**Supplementary Results**

**Figure S2. Scatter plot of Biological (BA) vs Chronological Age (CA) in the test set (N=4,772).**

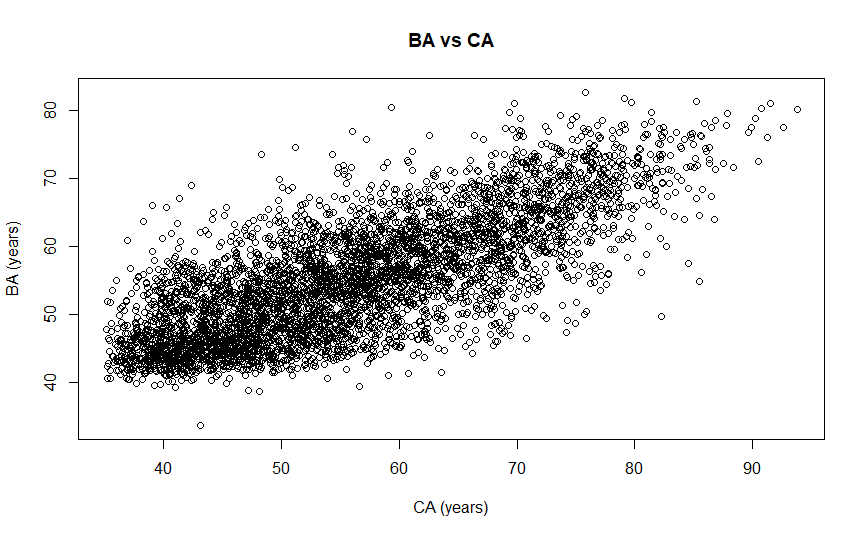

**Figure S3. Histogram of Δage values in the test set (N=4,772).**

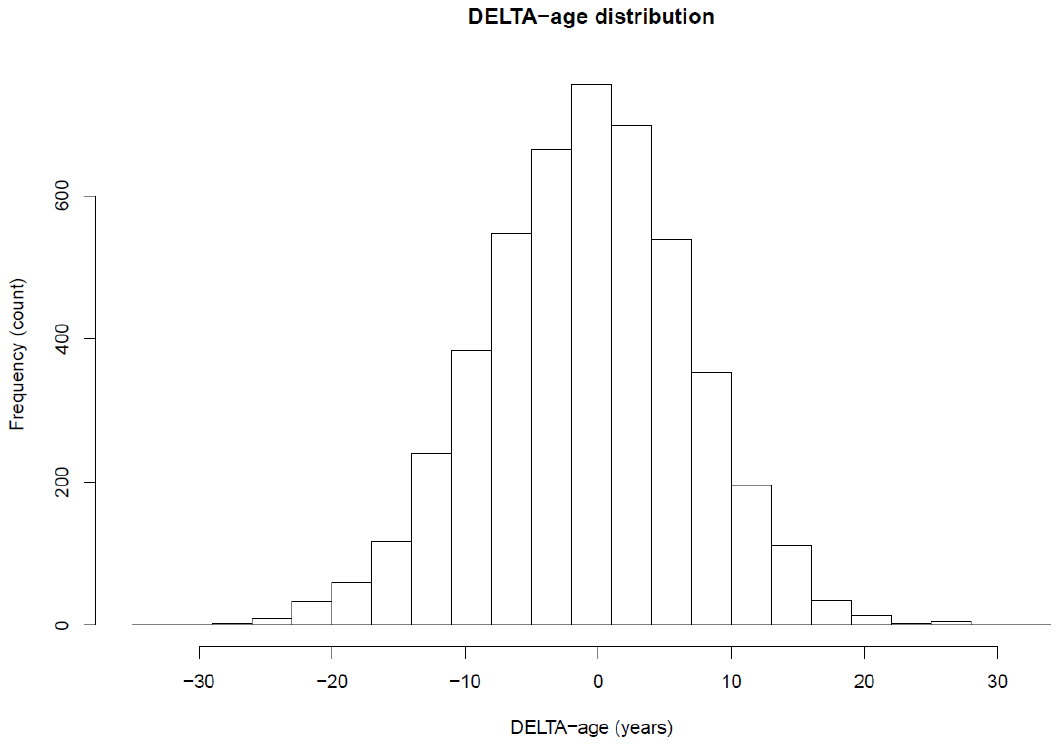

**Figure S4. Scatter plot of Δage vs Chronological Age (CA) in the test set (N=4,772).**

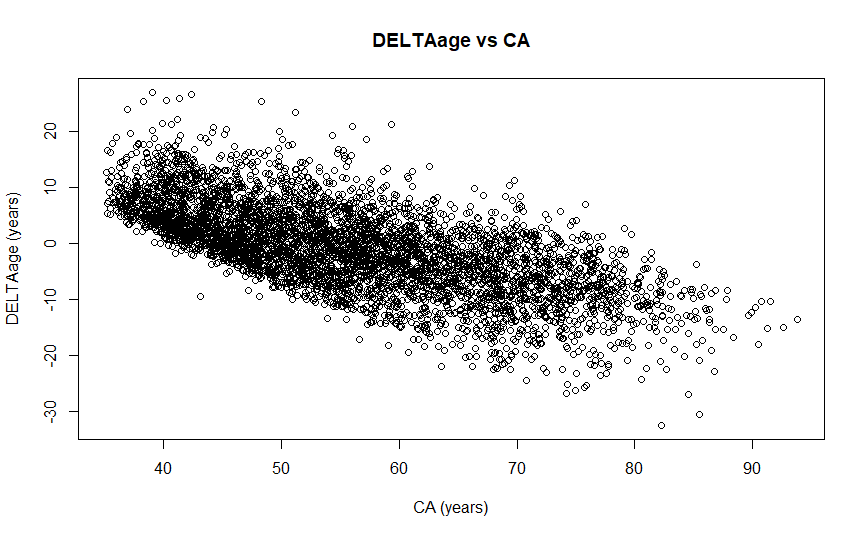

**Table S3. Association of Δage (BA-CA) with physical and mental wellbeing.**

| Wellbeing scale | Model 1  Beta (SE)  (p-value) | Model 2  Beta (SE)  (p-value) | Model 3  Beta (SE)  (p-value) | Mean (SD) |
| --- | --- | --- | --- | --- |
| SF-36 mental | **-0.30 (0.09)**  **(1.6×10^-3^)** | **-0.23 (0.09)**  **(0.01)** | -0.12 (0.09)  (0.22) | 46.93 (10.1) |
| SF-36 physical | **-0.81 (0.10)**  **(<2.0×10^-16^)** | **-0.67 (0.10)**  **(9.8×10^-12^)** | **-0.44 (0.10)**  **(1.8×10^-5^)** | 46.63 (6.4) |

All associations were incrementally adjusted for i) Model 1: sex and CA; ii) Model 2: Model 1 + prevalent health conditions (cardiovascular disease, diabetes, cancer); iii) Model 3: Model 2 + lifestyles/socioeconomic factors. Since these scales were computed from the same test (SF-36)[4], they were modelled together in a multivariable regression model, when available in the test set (N=3,728). Beta values indicate the variation in Δage (years) per Standard Deviation (SD) increase in the scale. Significant associations surviving correction for multiple testing (p < 0.025) are highlighted in bold.

**Table S4. Associations of Δage (BA-CA) with main lifestyles and socioeconomic factors in a stepwise multivariable regression model.**

| Lifestyle variable | Levels/Variables | Beta | SE | p | Class of exposures |
| --- | --- | --- | --- | --- | --- |
| **Smoking status** | Never smokers (reference) | - | - | - | Lifestyles  (or proxies) |
|  | **Current smokers** | **1.62** | **0.22** | **1.8×10^-13^** |  |
|  | Previous smokers | 0.58 | 0.21 | 6.6×10^-3^ |  |
| **Obesity (BMI)** | <25 Kg/m2 (reference) | - | - | - |  |
|  | [25-30[ Kg/m2 | 0.14 | 0.21 | 0.50 |  |
|  | **≥ 30 Kg/m2** | **1.67** | **0.23** | **1.8×10^-13^** |  |
| Adherence Score  to Mediterranean Diet [8] | Low (0-3) | - | - | - |  |
|  | Moderate (4-6) | -0.29 | 0.19 | 0.13 |  |
|  | High (7-9) | -0.59 | 0.31 | 0.06 |  |
| Alcohol consumption | Occasional/current drinkers  (1–12 g/day) (reference) | - | - | - |  |
|  | Life-time abstainers | 0.64 | 0.23 | 6.1×10^-3^ |  |
|  | Former drinkers | -0.10 | 0.42 | 0.81 |  |
|  | Current drinkers (24.1-48 g/day) | 0.07 | 0.27 | 0.79 |  |
|  | Current drinkers (12.1-24 g/day) | 0.22 | 0.27 | 0.42 |  |
|  | Current drinkers (>48 g/day) | -0.54 | 0.33 | 0.10 |  |
| **Occupational class** | Professional and managerial (reference) | - | - | - | SES |
|  | Non-manual skilled | 0.08 | 0.23 | 0.72 |  |
|  | Manual unskilled | 0.37 | 0.28 | 0.20 |  |
|  | Manual skilled | 0.68 | 0.28 | 0.02 |  |
|  | Retired/housewife | 1.18 | 0.44 | 8.1×10^-3^ |  |
|  | **Unemployed/unclassified** | **2.23** | **0.76** | **3.5×10^-3^** |  |
| Yearly household income  (Euros) | <10.000  (reference) | - | - | - |  |
|  | [10.000-25.000] | 0.38 | 0.37 | 0.30 |  |
|  | ]25.000-40.000] | -0.47 | 0.40 | 0.24 |  |
|  | >40.000 | -1.02 | 0.44 | 0.02 |  |
|  | Not declared | -0.19 | 0.37 | 0.61 |  |
| Housing | Rented flat (reference) | - | - | - |  |
|  | Dwelling ownership | 0.52 | 0.29 | 0.07 |  |
|  | >1 dwelling ownership | 0.08 | 0.40 | 0.84 |  |

Significant associations surviving correction for multiple testing (p < 5×10^-3^) are highlighted in bold. All lifestyles and socioeconomic (SES) variables not shown here were associated with an AIC increase and were therefore excluded by the stepwise regression procedure.

**Moli-sani Study Investigators**

The enrolment phase of the Moli-sani Study was conducted at the Research Laboratories of the Catholic University in Campobasso (Italy), the follow up of the Moli-sani cohort is being conducted at the Department of Epidemiology and Prevention of the IRCCS Neuromed, Pozzilli, Italy.

**Steering Committee:** Licia Iacoviello*°(Chairperson), Giovanni de Gaetano* and Maria Benedetta Donati*.

**Scientific secretariat:** Licia Iacoviello*° (Coordinator), Marialaura Bonaccio*, Americo Bonanni*, Chiara Cerletti*, Simona Costanzo*, Amalia De Curtis*, Giovanni de Gaetano*, Augusto Di Castelnuovo^§^, Maria Benedetta Donati*, Francesco Gianfagna°^§^, Mariarosaria Persichillo*, Teresa Di Prospero* (Secretary).

**Safety and Ethycal Committee:** Jos Vermylen (Catholic Univesity, Leuven, Belgio) (Chairperson), Ignacio De Paula Carrasco (Accademia Pontificia Pro Vita, Roma, Italy), Antonio Spagnuolo (Catholic University, Roma, Italy).

**External Event adjudicating Committee**: Deodato Assanelli (Brescia, Italy), Vincenzo Centritto (Campobasso, Italy).

**Baseline and Follow-up data management:** Simona Costanzo* (Coordinator), Marco Olivieri (Università del Molise, Campobasso, Italy).

**Informatics:** Marco Olivieri (Università del Molise, Campobasso, Italy).

**Data Analysis:** Augusto Di Castelnuovo^§^ (Coordinator), Marialaura Bonaccio*, Simona Costanzo*, Alessandro Gialluisi*, Francesco Gianfagna*^§^, Emilia Ruggiero*.

**Biobank and biomedical analyses:** Amalia De Curtis* (Coordinator), Sara Magnacca^§^.

**Genetic analyses:** Benedetta Izzi* (Coordinator), Francesco Gianfagna°^§^, Annalisa Marotta*, Fabrizia Noro*.

**Communication and Press Office:** Americo Bonanni* (Coordinator), Francesca De Lucia (Associazione Cuore Sano, Campobasso, Italy).

**Recruitment staff:** Mariarosaria Persichillo* (Coordinator), Francesca Bracone*, Francesca De Lucia (Associazione Cuore Sano, Campobasso, Italy), Simona Esposito*, Cristiana Mignogna*, Teresa Panzera*, Livia Rago*, Emilia Ruggiero*.

**Follow-up Event adjudication:** Livia Rago* (Coordinator), Simona Costanzo*, Amalia De Curtis*, Licia Iacoviello*°, Teresa Panzera*, Mariarosaria Persichillo*.

**Regional Health Institutions:** Direzione Generale per la Salute - Regione Molise; Azienda Sanitaria Regionale del Molise (ASReM, Italy); Molise Dati Spa (Campobasso, Italy); Offices of vital statistics of the Molise region.

**Hospitals:** Presidi Ospedalieri ASReM: Ospedale A. Cardarelli – Campobasso, Ospedale F. Veneziale – Isernia, Ospedale San Timoteo - Termoli (CB), Ospedale Ss. Rosario - Venafro (IS), Ospedale Vietri – Larino (CB), Ospedale San Francesco Caracciolo - Agnone (IS); Casa di Cura Villa Maria - Campobasso; Fondazione di Ricerca e Cura Giovanni Paolo II - Campobasso; IRCCS Neuromed - Pozzilli (IS).

***Department of Epidemiology and Prevention, IRCCS Neuromed, Pozzilli, Italy

°Department of Medicine and Surgery, University of Insubria, Varese, Italy

^§^Mediterranea, Cardiocentro, Napoli, Italy

*Baseline Recruitment staff is available at*

[*http://www.moli-sani.org/index.php?option=com_content&task=view&id=21128&Itemid=118*](http://www.moli-sani.org/index.php?option=com_content&task=view&id=21128&Itemid=118)
